# Accuracy of Smartphone-based Vital Monitoring Using Remote Photoplethysmography Technology Enabled WellFie application

**DOI:** 10.1101/2023.01.14.23284548

**Authors:** Sujata Rajan, Madhava Sai Sivapuram, Shiv Shankar Kumar, Vivek Podder

## Abstract

**Background:** Remote health monitoring technologies gained interest in the context of COVID-19 pandemic with potential for contactless monitoring of clinical patient status. Here, we examined whether vital parameters can be determined in a contactless manner using a novel smartphone-based technology called remote photoplethysmography (rPPG) and compared with comparable certified medical devices.

**Methods:** We enrolled a total of 150 normotensive adults in this comparative cross-sectional validation study. We used an advanced machine learning algorithm in the WellFie application to create computational models that predict reference systolic, diastolic blood pressure (BP), heart rate (HR), and respiratory rate (RR) from facial blood flow data. This study compared the predictive accuracy of smartphone-based, rPPG-enabled WellFie application with comparable certified medical devices.

**Results:** When compared with reference standards, on average our models predicted systolic blood pressure (BP) with an accuracy of 93.94%, diastolic BP with an accuracy of 92.95%, HR with an accuracy of 97.34%, RR with accuracy of 84.44%. For the WellFie application, the relative mean absolute percentage error (RMAPE) for HR was 2.66%, for RR was 15.66%, for systolic BP was 6.06%, and for diastolic BP was 7.05%.

**Conclusion:** Our results on normotensive adults demonstrates that rPPG technology-enabled Wellfie application can determine BP, HR, RR in normotensive participants with an accuracy that is comparable to clinical standards. WellFie smartphone application based on rPPG technology offers a convenient contactless video-based remote solution that could be used in any modern smartphone.

## Introduction

The measurement of physiological parameters including heart rate, respiratory rate, blood pressure is fundamental to assess human health. Modern biotransformation solutions enabled low cost, non-invasive hospital-based health monitoring systems.[1] However, the physiological parameters are believed to be best analyzed in the patient’s environment. Typically, in clinical practice these parameters are measured using contact-based devices which are not optimized to capture in the patient’s environment. In addition, they pose some intrinsic limitations such as loss of contact, skin irritations in patients with fragile skin compromising the reliability of measurements and cannot be used for long-term follow-up or continuous remote monitoring. [2,3] Lack of remote monitoring medical devices is a major limitation to overcome those barriers. However, to overcome these barriers contactless and remote health monitoring technologies are gaining much interest in the context of the global COVID-19 pandemic. [4] Camera-based remote photoplethysmography (rPPG) technology is such a novel method that offers contactless remote vital signs monitoring and that detect pulse-induced subtle color variations from the human skin using a multi-wave Red, Green, Blue (RGB) camera. These technologies have numerous fields of application where it is suitable, reliable, accurate, and advantageous ranging from clinical and telemedicine setting to occupational setting, sports science, automotive field to emotion recognition. [5, 6, 7, 8,9] In inpatient or ICU, contactless remote vital monitoring enables a protective environment for both patients and physicians while also enabling healthcare team to increase attention to patient conditions.[10] In telehealth, remote monitoring of vital signs play a critical role in the assessment and diagnosis. [11] For example, sepsis is a dreadful condition of a variety of infectious diseases where vital parameters play a critical role in making a management decision. Similarly, for a range of non-communicable diseases such as hypertension, cardiac arrhythmias, chronic respiratory conditions, mental illnesses, frequent and long-term follow-up with vital parameters is required to optimize their treatment plans. [12] As such, remote technological solutions can increase healthcare accessibility to remote places and low-income families and remote access to variety care services including child care, chronic disease care, psychiatry care. [12]

For example, hypertension is a major global public health problem affecting more than 25% of the adult population worldwide.[13] Globally, 1.3 billion adult population are estimated to have hypertension and disproportionately two-third of them living in low- and middle-income countries. [13] Hypertension also is a major risk factor for cardiovascular diseases, cerebrovascular diseases, chronic kidney disease, dementia, and major cause of premature death globally. [13,14] The World Health Assembly (WHA) has adopted a global target of 33% reduction in hypertension between 2010 and 2030 to fight with rapidly growing non-communicable diseases (NCDs).[15] Epidemiologic studies reported that 46% of the adult hypertensive population are undiagnosed and only 21% of those diagnosed have a hypertension control. [16]

Clinical practice guidelines recommended early detection of hypertension by screening high-risk target groups which can result in prompt treatment with reduction in related mortality and morbidity and health care costs.[17] However, screening for diagnosing and assessing the response to therapy for hypertension are subjected to variability of current blood pressure measurement techniques. Different potential measurement conditions throughout daily life can be responsible for the variability of the measurement methods. For example, conventional blood pressure measurement methods are inconvenient and require inflatable cuff-based technology. Moreover, due to masked hypertension (MHT) including masked uncontrolled hypertension (MUCH), defined as BP in the hypertensive range when measured outside of the clinic.

To fight this epidemic, many recent hypertension guidelines have highly recommended non-clinic blood pressure (BP) monitoring utilizing standard automated BP monitors and ambulatory BP monitors. [17] It gives patients and physicians representative BP trends throughout the day. However, standard automated blood pressure machines are not convenient to use outside the home, not widely available, not standardized for accuracy, and cost ineffective. Whereas ambulatory BP monitoring are inconvenient to wear throughout the day. [18,19] Therefore, a better technology is required that offer comparable accuracy, comfortable and convenient measurements anywhere and anytime without causing a financial burden.

A novel smartphone-based rPPG technology can meet these requirements. The rPPG technology is a camera-based solution for non-contact cardiovascular monitoring, proven to be as accurate as traditional PPG devices. [20] This is a technology of contactless recording of a PPG signal with a Red-Green-Blue (RGB) camera. This technology for imaging blood flow patterns from facial video capture, can process imperceptible facial blood flow oscillations giving comparable results with the help of advanced machine learning models. [20,21] Contact PPG devices may not be suitable for vital sign monitoring during contagious conditions like coronavirus disease 2019 (COVID-19). In such scenarios, contactless rPPGs can enable assessing patients requiring non-contact approaches or cannot physically consult a physician. In addition, with a growing global adoption of smartphone devices, rPPG technology can be integrated into those devices using mobile applications particularly in resource-constrained settings. Previous studies showed proof of validity in monitoring for different cardiac and respiratory parameters (blood pressure [BP], heart rate [HR], respiratory rate [RR]) by using this technology.[20,21] A recent systematic review and meta-analysis showed that consumer-grade contactless vital sign monitors are accurate in measuring HR compared to medical devices and a need for large clinical validation studies that is conducted in clinical settings and included other vital parameters.[22] In another hospital trial, rPPG technology was found to have a good reliability for RR estimation in real-life clinical settings. [23]

In this study, thus, we hypothesize that signals extracted from facial blood flow oscillations can indicate heart rate, respiratory rate, systolic and diastolic BP. We examined whether blood pressure, heart rate, respiratory rate can be measured using a contactless smartphone-based novel rPPG technology which processes facial blood oscillations from facial video captured with a camera and advanced machine learning to determine the parameters.

The objective of the study is to measure and determine clinical accuracy of key health vitals output from WellFie contactless smartphone-based novel technology using facial video capture in comparison to comparable certified medical devices.

## Methods and Materials

### Study Design

A comparative cross-sectional validation study was conducted between 20^th^ April 2022 to 26^th^ May 2022 at the Ayurvedic and Unani Tibbia College and Hospital, Government of National Capital Territory of Delhi, New Delhi, India. The study investigated the comparative clinical accuracy between Wellfie application solution using remote photoplethysmography technology (rPPG) and certified medical devices.

### Subject Selection, Enrollment, and informed Consent

The participants were recruited between the age group of 18 years and 70 years. The participants were enrolled after obtaining a proper written informed consent. The process of obtaining informed consent was in accordance with applicable regulatory requirement (s) and adhered to good clinical practice (GCP) and to the ethical principles. The participants were adequately explained about the study. The demographic details of the participants along with clinical history were captured from 150 participants using a convenience sampling method throughout the study period. The study investigators assessed the eligibility criteria after data collection and enrolled a total of 150 participants within a 36-day duration. Any known medical history of the enrolled participants were recorded.

#### Inclusion Criteria

a. Participant’s age group between 18 – 70 years.
b. Healthy participants with normotensive BP range (systolic ≥ 100 to < 140 mmHg and diastolic ≥ 60 to < 90 mmHg)
c. Participants should sign the informed consent and agree for audio and video recording of the entire procedure.

#### Exclusion Criteria

a. Participants with cancer, epilepsy, psychological disorders, and other severe illness
b. Participants with deep cuts and scars in face

### Study Procedures

All the enrolled participants were rested in a sitting position and monitored for non-invasive vital signs (blood pressure, heart rate, respiratory rate) simultaneously using both the certified medical devices and the WellFie application. Blood pressure cuff was tied to the left arm and the pulse oximeter sensor was put on the right index finger (for respiratory rate estimation) to monitor the vital signs (certified medical devices were used). At the same time, the participants were asked to face in front of a camera for 60 seconds to capture the vital signs (WellFie application with rPPG system was used). Both procedures were audio and video recorded. The data were manually noted into excel sheets for a statistical analysis. The detail steps of the study procedures are following:

1. Participants were ensured to take 15-minute complete rest, to avoid consumption of any beverages (coffee/tea/soft drinks) and smoking within the past 30 minutes of the test.
2. Any comorbidities were recorded before starting the test
3. Participants were asked to sit resting in a chair and then camera-based Wellfie application + comparison medical devices were set up
4. During the test period, the WellFie application and the comparison medical devices were ensured to run at the same time
5. Each test was taken for the entire 60 seconds period till WellFie application and medical device completes the test
6. Each test was repeated times in intervals of 2-3 minutes and only the last measurement was considered for the analysis.

### Data Collection and Processing

This study utilized a case report form (CRF) to record data. The principal research investigator ensured the accuracy and completeness of the recorded data and provided signatures on appropriate CRFs and then documented them in compliance with local regulations. Visual and/or computer data was performed to identify possible data discrepancies. The site staff was responsible for resolving all queries in the database.

### Outcome Assessment

The clinical accuracy of key health vitals output (BP, heart rate, respiratory rate) from WellFie application in comparison to certified medical devices was assessed

### WellFie Remote Photoplethysmography Technology (rPPG)

The WellFie application has a built-in Remote Photoplethysmography (rPPG) technology and an integrated algorithm that offers a camera-based solution for accurate non-contact cardiovascular monitoring (heart rate, respiratory rate, blood pressure) compared to traditional PPG devices. This technology measured the changes in red, green, and blue light reflection from the human skin using a multi-wave RGB camera (a camera equipped with CMOS sensor that delivers colored images of persons and objects by capturing light in red, green, and blue wavelengths [RGB]), quantified the contrast between specular reflection and diffused reflection. WellFie extracted vital signs using a signal taken from the upper cheek region of the face and analyzed it with advanced AI and signal processing. The entire functionality of this technology is illustrated in figure 1.

**Figure 1:**
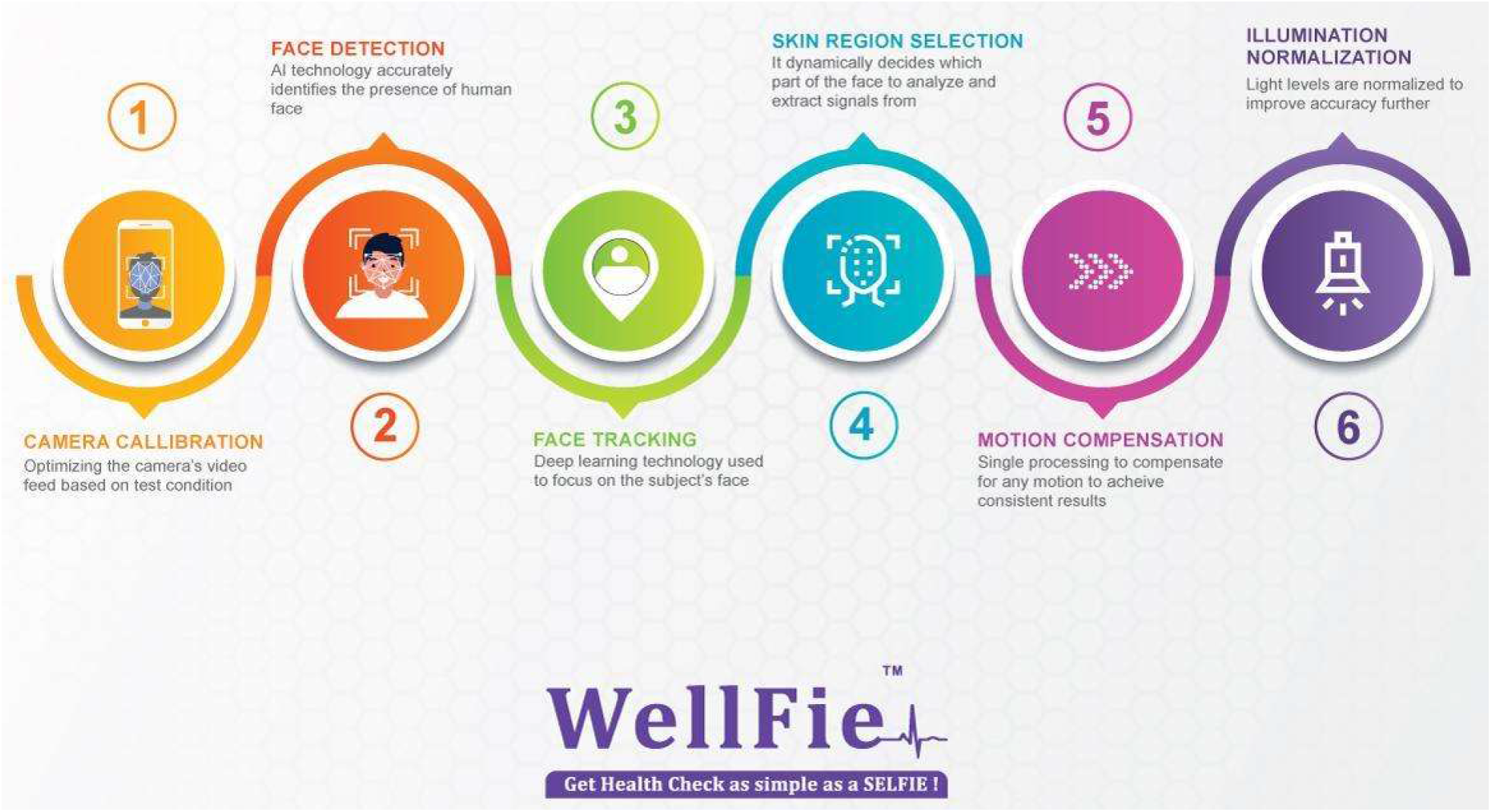

### Technology Requirements

Wellfie application can be used in any smartphone/tablet/computer/laptop with an android/IOS operating system and web camera (inbuilt or external). The operating system of the camera can be used only if it can produce at least 20 frames per second to run Wellfie’s application. For this study, we have used an external Logitech C920 HD pro web camera which was attached to a laptop (hp - 11th Gen Intel® Core™ i5 processor) and can produce 30 frames per second. The certified medical devices used for vital monitoring were: Pulse Oximeter (Masimo Mightysat SatRx) and Blood Pressure Device (Omron). The luminosity of the study environment was pre-checked using the lux meter app to ensure a minimum of 300 lux in the room.

### Confidentiality

All participant identification records were kept confidential as per the applicable laws and regulations. Only participant numbers and initials were recorded in CRF. Serial number was appointed (eg, AAA, BBB) in cases where local law restricts using participant initials. The study findings were securely stored in the computer in accordance with local data protection laws. The participants were instructed that sponsoring or regulatory authorities may inspect their medical and personal records for verification with strict precaution in accordance with local data protection laws. The participants were informed that study details without participant identifiers will be posted on the clinical trial registry website.

### Monitoring Procedures

Study sites were visited by an authorized WellFie application solution representative for inspection of study data, participants’ medical records, CRFs in accordance with current ICH-GCP guidelines and respective local and national government regulations and guidelines as applicable. Monitoring was performed in person or remotely.

### Statistical Analysis

There are four models developed to predict heart rate (HR), respiratory rate (RR), systolic and blood diastolic blood pressure, respectively. To validate the prediction accuracy of the model real data of 150 individuals are collected and further predicted values are collected by running all four models. The predictive models were validated considering the accompanying statistical measures:

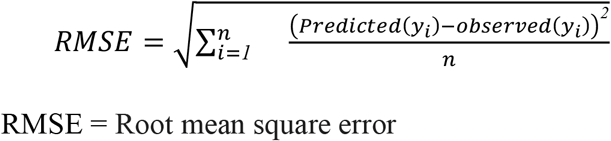

To check the predictive accuracy of each model relative mean absolute percentage error (RMAPE) was also used. The RMAPE is widely used to validate predictive accuracy, which provides an indication of the average size of prediction error expressed as a percentage of the relevant observed value.

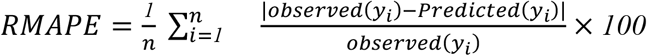

and Further

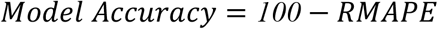

### Statement of Compliance

This clinical investigation was conducted in accordance with the ethical principles that have their origin in the Declaration of Helsinki, ISO 14155:2011 (Clinical Investigation of Medical Devices for Human Subjects – Good Clinical Practice), ICH-GCP, and any regional or national regulations, as appropriate.

### Ethical Clearance Statement

The study was conducted after getting the ethical clearance from the Institutional Ethics Committee (IEC) of the Ayurvedic and Unani Tibbia College and Hospital vide IEC reference no: No.F.5(283)2013-COPF(Ayurveda)/672 (dated 21/12/21). The clinical trial registration number for this study is CTRI/2022/01/039530.

## Results

We recruited 150 participants whose characteristics are listed in table 1. In this study, total participants constituted both female (65.4%) and male (34.6%) participants. 46.7% of The total participants represented underweight (8%), normal weight (48%), and overweight (46.7%). As shown in table 2, high rates of prediction accuracy in terms of predicting the reference BPs, RR, HR were observed by all models in the independent dataset of our validation cohort. Mann-Whitney U test was also conducted in order to further guarantee the high rate of accuracy. The goal of these two tests is to determine whether the observed and predicted values have similar or different trends. All models are about to predict results with a similar trend as they had found in the machine data. The complete results are displayed in table 1 and the distribution shape and trend of the actual and predicted values are illustrated in Figure 2.

**Table 1:** Participant Characteristics

| Participant Characteristics | Number (n=150) | Percentage (%) |
| --- | --- | --- |
| **Gender** |  |  |
| Male | 52 | 34.6% |
| Female | 98 | 65.4 |
| **Age** |  |  |
| 18-29 | 64 | 42.7% |
| 30-49 | 49 | 32.7% |
| 50-64 | 31 | 20.7% |
| 65+ | 6 | 4% |
| **Body Mass Index** |  |  |
| < 18.5 | 8 | 5.3% |
| 18.5 - 24.9 | 72 | 48% |
| > 24.9 | 70 | 46.7% |

**Table 2:**
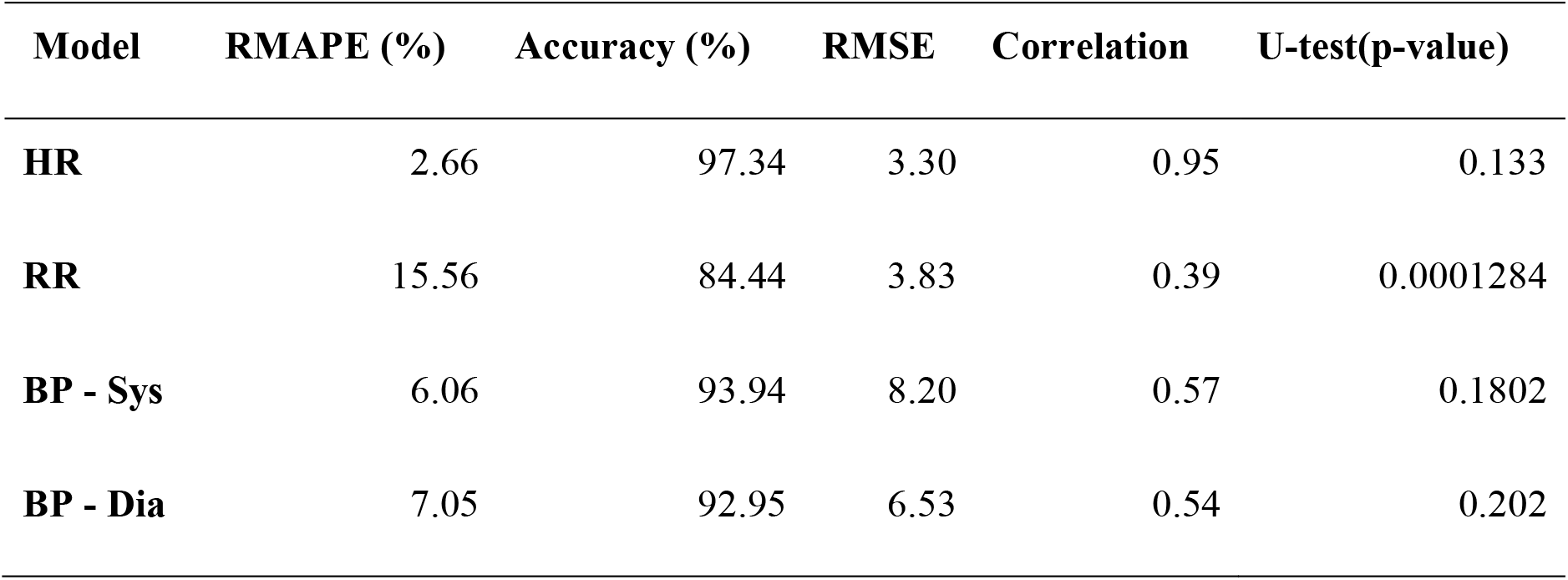
Model prediction performance evaluation.

**Figure 2:**
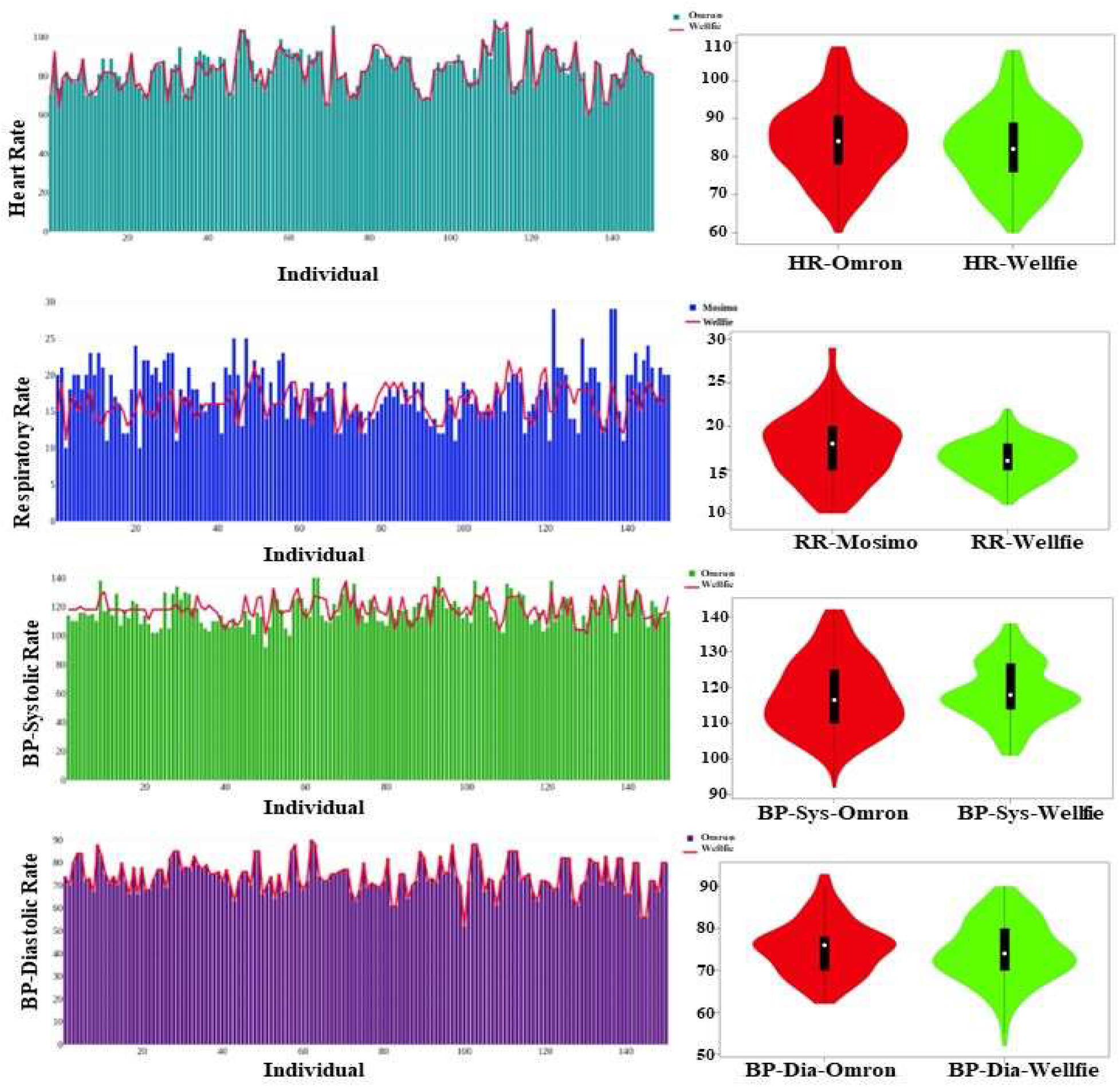

### Remote photoplethysmography Determines BPs, HR, and RR

When compared with reference standards, on average our models predicted systolic BP with an accuracy of 93.94%, diastolic BP with an accuracy of 92.95%, HR with an accuracy of 97.34%, RR with accuracy of 84.44%. The Mann-Whitney (U) tests for predictive accuracy were for systolic BP (0.18), diastolic BP (0.20), HR (0.13), and RR (0.0001). Our findings corresponded to correlations of 0.57, 0.54, 0.39, and 0.95 for systolic BP, diastolic BP, RR, HR, respectively.

### Percent Error

As per the recommendations from the Association for the Advancement of Medical Instrumentation, the Consumer Technology Association and the American National Standards Institute, the acceptable error rate for a physical monitoring device is <±10%. [24] In our study, the RMAPE was used to determine the acceptable error rate and thereby, to validate the predictive accuracy. For the WellFie application, the RMAPE for HR was 2.66%, for RR was 15.66%, for systolic BP was 6.06%, and for diastolic BP was 7.05%.

## Discussion

During COVID-19 pandemic, the contactless vital monitoring technologies has gained increased attention among researchers and physicians.[26,27,28] They can measure vital signs by analyzing video-recorded through digital cameras.[29] Video-based rPPG technology can extract the color changes in light reflection from human facial skin and then use them to accurately predict BP, RR, and HR.[30] Our study revealed a high predictive accuracy in the monitoring of cardiac and respiratory (BP, HR, RR) parameters by using WellFie smartphone-based rPPG application when compared to well-based FDA-approved medical devices. We observed that rPPG signal obtained in the face closely corresponds to BP, HR, RR wave oscillations obtained simultaneously in the finger and left arm using reference device technologies. We determined the predicted error by measuring the RMAPE for systolic BP (6.06%), diastolic BP (7.05%), RR (15.56%), and HR (2.66%). Thus, the present study demonstrates that rPPG technology can determine BP, HR, RR in normotensive participants with an accuracy that is comparable to clinical standards. For the currently available BP monitors, a consensus report from Association for the Advancement of Medical Instrumentation/European Society of Hypertension/International Organization for Standardization (AAMI/ESH/ISO) considered a BP medical device acceptable if its estimated probability of a tolerable error (≤10 mmHg) is at least 85%. [31] However, true validation will require testing on non-normotensive participants according to the methodology outlined in this standard. Our findings indicate that blood flow through facial vasculature has useful information about BP, HR, RR as compared to FDA-cleared BP and pulse oximetry devices.

Video-based contactless vital monitoring offers several advantages over other contact-based monitoring technologies in different settings.[31-35] For example, contactless methods can account for the naturally occurring and cyclic BP fluctuations through short continuous measurements, do not require a cuff making it comfortable, and easily implementable, high adaptability, scalability through any smartphone device without a need for any special equipment.[30,27] These advantages enable this technology to be used for remote personal health status assessment, clinical follow-up, integrated clinical exams, remote care of elderly and patients with chronic illness, immobility, recovering at home. These will further add to reducing the strained healthcare system and improved mortality and morbidity due to inaccessible and prompt healthcare support.

As the need for virtual care grew with a high adaptability rate, the artificial intelligence (AI) is transforming telemedicine care across the globe. [31-39] WellFie rPPG technology with high predictive accuracy can offer an integral link whereby a virtual physician can gain real-time contactless insights on patient clinical data. A systematic review evaluating camera-based vital monitoring found that RGB cameras can reliably monitor HR and RR in a controlled environment [20]; while our study had a comparatively low predictive accuracy for RR, we found a very high predictive accuracy for both systolic and diastolic BP and HR. However, investigation in real-world settings might have shown a different accuracy.

### Limitations

Despite the high predictive accuracy of rPPG for determining the cardiac and respiratory parameters, further investigation is warranted. For example, in our study we included participants in the normotensive BP ranges without investigating on hypertensive or hypotensive BPs or with comorbid conditions who also might be taking medications. Therefore, future studies should recruit hypertensive or hypotensive participants and whether they are taking medication or not to test the predictive accuracy of current normotensive-based computation models. In our study, computational BP models were validated against reference BP values from certified BP and pulse oximeter devices. While certified medical devices were validated against gold standard techniques (e.g., invasive intra-arterial or a mercury sphygmomanometer); our technology was not validated against the gold standards warranting further investigation. To further test the capabilities of the technology, future studies should also control for different conditions including outdoors, in moving vehicles, low light conditions, etc.

### Potential Applications and Future Research

Despite the limitations of the study, our study showed that WellFie’s smartphone application based on rPPG technology offers a convenient contactless video-based remote solution that could be used in any modern smartphone.[40] Our investigated smartphone application with rPPG technology would be comfortable, adoptable in remote location compared to cuff-based devices, allowing for BP measurements at a condition that can potentially prevent white-coat and masked hypertension effects and also can be applied in variety of health settings ranging from intensive care units, telehealth, home monitoring, sports exercise, etc. In our study, we measured vital signs in resting individuals as clinical measurements are taken under resting conditions. However, future research can investigate whether it can track BP while physical exercise, opening a new avenue of research for the science of fitness tracking devices, exercise, and cardiac stress tests where traditional methods of BP measurements have been challenging. The wide-validation initiatives will yield accurate, convenient, widely available BP measurement technologies in smart devices. These will translate into frequent capture of BP and HR measurements in the population both in research and clinical care facilitating a better arena for hypertension and other cardiorespiratory disorders management.

## Data Availability

All data produced in the present study are available upon reasonable request to the authors.

## Acknowledgement

The authors would like to express their gratitude to BWell HealthTech Pvt Ltd for their logistical support.

